# Vaccination of adolescent girls against Human Papillomavirus (HPV) in Embakasi West, Kenya: the role of family ties and value systems in decision making

**DOI:** 10.64898/2025.12.01.25341402

**Authors:** Wanjiru S. Ng’ang’a, Solomon K. Cheboi, Peter Omemo, Apollo O. Maima

## Abstract

**Background and Objective:** Uptake of the HPV vaccine among adolescent girls in Embakasi West sub-County, Kenya has lagged behind since the introduction of the vaccine in 2019. While family, friends and community form a strong and influential support system for individuals, their influence on HPV vaccine uptake is undocumented.

**Materials and Methods:** A mixed methods study was undertaken in Embakasi West Sub-County, Nairobi, Kenya in which 371 school-going girls responded to a self-administered questionnaire, along with three caregiver and two adolescent focus group discussions, and 13 key informant interviews. The influence of family, peers, religion and culture on uptake of the HPV vaccine were examined. Quantitative data were analysed using descriptive and inferential statistical analysis while qualitative data were thematically analysed.

**Results:** Majority (92.2%) of respondents had siblings, 74.4% resided with both parents, and 30.7% identified as Pentecostal Christians. Half (50.1%) of respondents reported their parents wanted them to be vaccinated, 79.2% could discuss HPV with their parents but 41.5% had not had such discussions. Overall, 20.8% of respondents had an HPV-vaccinated relative while 19.8% had received negative information about the vaccine. Religious and cultural acceptability of the vaccine was at 52.8% and 48.2% respectively. Parent attitude towards adolescent vaccination, (p<0.001), discussing HPV in the home (p<0.001), having a vaccinated family member (p=0.01), positive attitude of peers (p=0.02), beliefs about culture (p<0.001) and religion (p<0.001) were significantly associated with vaccine uptake. However, limited HPV-related discussion in the home negatively affected uptake (OR 0.230 95% CI [0.098-0.541], *p*=0.001). Caregiver knowledge and understanding were seen as means to overcome family opposition, religion and cultural beliefs.

**Conclusion:** Substantial consideration is given to family values in the medical decision-making process of caregivers for their children. Interventions that focus on enhancing caregiver-child dialogue could be used to drive uptake of the HPV vaccine.

## Introduction

Comprising of more than 200 strains, the Human Papillomavirus (HPV) is the most common sexually transmitted infection in the world and can be classified as either high or low risk. High risk strains are strongly linked to the development of several cancers. The majority of oropharyngeal and anal malignancies in both men and women, as well as vaginal cancers are caused by high-risk HPV strains accounting for at least 690,000 new cases annually (1). Globally, cervical cancer has the highest incidence, prevalence and mortality of all cancers currently linked to HPV. Cervical cancer is the 8^th^ most common cancer globally, accounting for 3.3% of all cancer cases and ranked 9^th^ for all cancer mortality, causing 3.6% of all cancer-related deaths (2). Based on the current trajectory, the number of new cervical cancer cases is projected to increase to over a million annually by the year 2050, up from an approximate 662,000 cases (3). Low-and middle-income regions of south America, south east Asia and Sub-Saharan Africa disproportionately carry the highest burden of cervical cancer with incidence and mortality estimates as high as 84% and 90% of the global cases respectively (3–6). A staggering 43% of global cervical cancer cases were reported in sub-Saharan Africa where Eastern Africa leads with an age standardized incidence rate (ASIR) of 40.42 and age-standardized mortality rate (ASMR) of 28.87 compared to global rates of 14.12 and 7.08 respectively (3). In Kenya, an estimated 5236 women were diagnosed with cervical cancer in 2023 with more than half that number succumbing to the disease, (7).

While cervical cancer has most often been diagnosed in women above the age of 50, there is an increasing number of new cases in women below the age of 50 (8). The implications of a diagnosis of cancer extend far beyond the individual’s health, spilling over to the economic and psycho-social and physical well-being of the family (9–11). Women under age 50 are not only of child bearing age, but often have young children in need of their care. As many as half a million children became maternal orphans having lost their mothers to breast or cervical cancer, and twice as many from all cancers (12). Late diagnosis, advanced disease and sub-optimal or inaccessible care result in higher mortality rates, as is the case in less developed nations, leading to familial and societal destabilization due to lost household income and support systems, compounding the effect of such cancers in the homes (12–15)

The HPV vaccine is a preventative vaccine administered to males and females between the ages of 9 and 26 for protection against selected HPV types. As a result of the burden of cervical cancer, the HPV vaccine was recently included in a 3-prong global initiative to eliminate cervical cancer as a public health concern. The initiative aims to vaccinate adolescent girls by their 15^th^ birthday, and prior to any exposure to the virus through sexual contact (16) effectively preventing future cases of cervical cancer in their adult lives. However, HPV vaccine coverage and uptake continues to remain below the 90% global goal due to challenges in access and hesitancy among caregivers responsible for consent (17). In 2023, 17% of age-eligible girls were vaccinated against HPV in Nairobi, against a national uptake of 33% in Kenya (18)

In most African countries, family, friends and community form a strong and influential support system for access and utilization of healthcare. (19). Driven by religion, extended family, interpretation of the illness, residential setting, relation to the head of household and cultural socialization, this support system may speak into health decisions including vaccine uptake. (20–23). While familial support has reportedly resulted in an increase in uptake of the HPV vaccine regionally (24), previous studies have not evaluated the association between the relational, contextual and social networks of school girls in Nairobi and uptake of the HPV vaccine. This study therefore examined the influence of social relations and cultural values on the uptake of the HPV vaccine by adolescent girls locally.

## Materials and Methods

### Study design and setting

A cross-sectional study entailing concurrent quantitative and qualitative approaches was carried out in Embakasi West Sub-County, Nairobi County-Kenya. The study was undertaken between July 24^th^ and October 18^th^, 2024 to assess the social and cultural values that influence uptake of the HPV vaccine among 10–14-year-old girls. The triangulation of both quantitative and qualitative data provided deeper understanding of feelings, experiences and perceptions of participants on the HPV vaccination program. The study was undertaken in Embakasi West, one of 17 Sub-Counties in Kenya’s capital city of Nairobi, where low levels of HPV vaccine uptake had been recorded repeatedly.

### Study population

The study population comprised of adolescent girls between the ages of 10 and 14 years, who attend public primary and junior secondary schools within the Sub-County, due to the large student population in these schools. In Kenya, the HPV vaccine is offered to all girls within this age group through school vaccination as well as at Health Facilities. Girls aged 10-14 years, and caregivers with adolescent girls in the target age group participated in the Focus Group Discussions (FGD). Health professionals of various cadres, community health workers and school staff with knowledge on the HPV vaccination program were interviewed as key informants.

### Sample size determination

The sample size was determined based on Cochran formula (1977) n=Z² P(1-P)/d² using a confidence interval of 1.96, proportion prevalence of 65.45% (25) and statistical significance set at 5%. Factoring a non-response rate of 10%, the calculated sample size came to 385 participants. Additionally, a total of five (5) FGD and 13 Key Informant Interviews (KII) were conducted, directed by content saturation.

### Sampling Procedure

To increase the equality of selection from the target population, multi-stage stratified sampling was employed to choose study participants, (Figure 1). At the first stage, six of 11 public primary and junior secondary schools were proportionally and randomly selected from the four cluster wards in the Sub County. Each grade between grade 5 and 8 was designated as a stratum. These grades were selected based on guidance from a school health officer on estimated age of students at each grade. Probability proportional to size (PPS) sampling technique was used to calculate the number of respondents to be sampled from selected schools, grade and stream. Strata to be sampled were selected at the second stage whereby larger strata (grade) had higher probability of being sampled. At the third stage, an equal number of respondents were sampled from each of the 10 strata (streams) which reduced the probability of the more populated grades being sampled. The overall result is that each student had the same probability of being sampled. Thirty-nine (39) female students were hence selected using simple random sampling from the selected grade and stream (Table 1).

**Fig 1:**
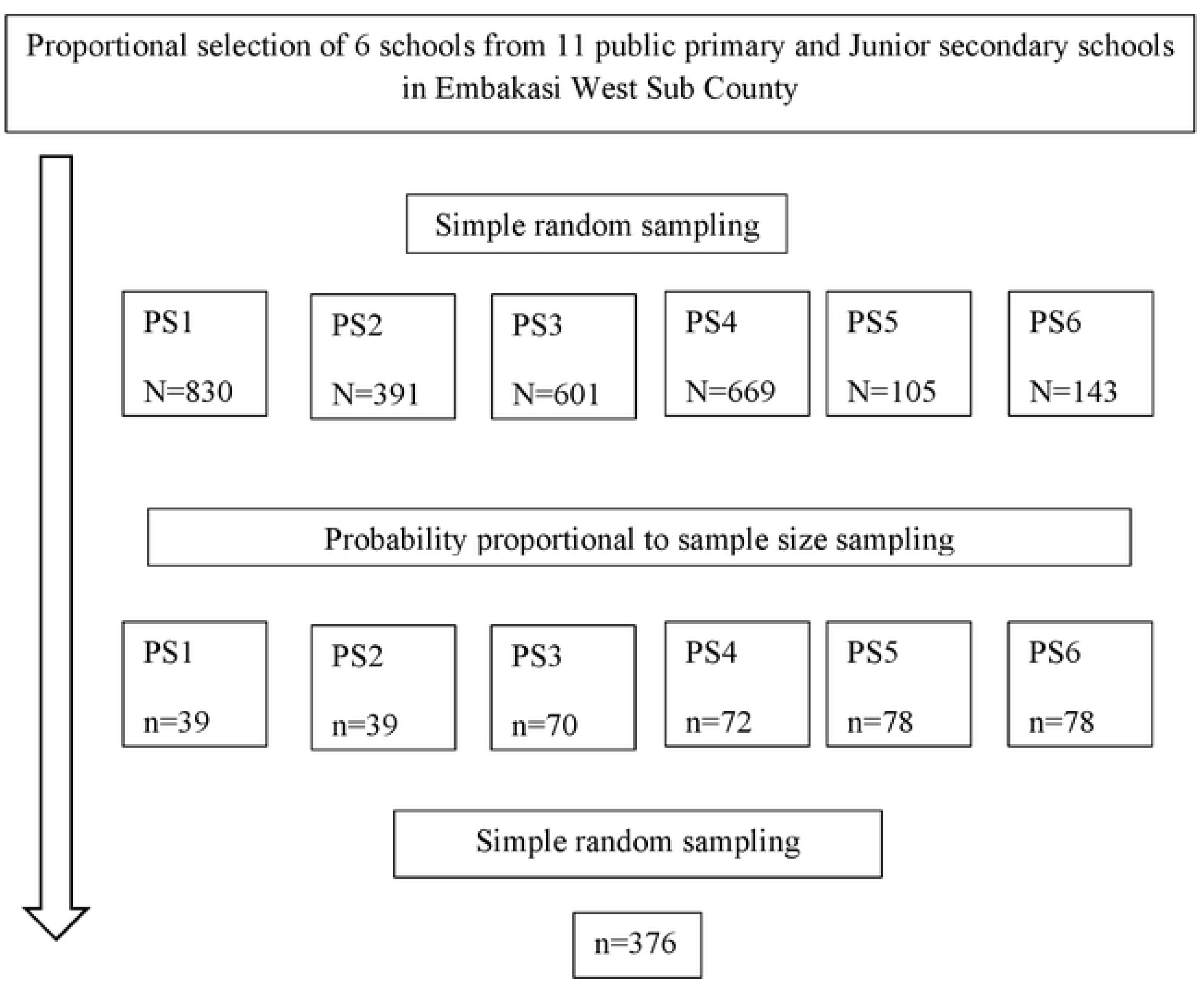
Schematic drawing of the sampling procedure for recruiting of participants attending school in Embakasi West Sub-County

**Table 1:** Multi-Stage Strata Sampling.

| Ward | # Of Schools In Ward | # Schools Selected | School | Grade | # Students per grade |
| --- | --- | --- | --- | --- | --- |
| Ward 1 | 1 | 1 | PS1 | G7 | 39 |
| Ward 2 | 5 | 2 | PS2 | G8 | 39 |
|  |  |  | PS3 | G5, G8 | 39 |
| Ward 3 | 4 | 2 | PS4 | G5, G8 | 39 |
|  |  |  | PS5 | G6, G8 | 39 |
| Ward 4 | 1 | 1 | PS6 | G6, G7 | 39 |

To enhance quality and uniformity in data collection, research assistants were trained on research ethics, interview tips, study objectives and methods prior to commencement of data collection. The survey tool was pre-tested and assessed for clarity and flow to ensure validity and reliability in a sub-sample of the target population and adjustments made to the tool. This site was excluded from actual data collection. The filled questionnaires were checked for completeness and data entry was done by the researcher for accuracy and confidentiality.

Qualitative data was collected from KII and FGD. Focus Group discussants were selected for participation in the separate adolescent and caregiver discussion groups. Girls were drawn from grades 7 and 8 in the selected schools, where two FGD were separately carried out at a secluded place within the schools. Women who resided within Embakasi West and had daughters aged 10 to 14 years were purposefully selected from 2 of the 4 wards in the Sub County, and invited to participate in three discussion groups held at health facilities in the ward. For the KII, interviewees were purposively selected from Facility Healthcare workers, Community health workers, and school staff, and each was interviewed in private at a convenient time and place.

### Data collection tools and techniques

A structured questionnaire was prepared in English, adapted from reviewing validated questionnaires in the literature from previous similar studies. The questionnaire was subjected to a readability test to ensure comprehension for the targeted age group, and also pre-tested in a small sub-sample of the study population. The survey tool had sections for assent; demographic information such as age, tribe, religion, birth order, sibling age, living with, and caregiver education level; familial, social and cultural aspects; and vaccine uptake defined as having received the 1^st^ and 2^nd^ dose. Data was collected using a self-administered questionnaire, supervised by the researcher and two assistants.

For qualitative data, an interview guide was prepared for the KII, and a discussion guide for the FGD. The guides had open-ended questions designed to encourage explanatory responses on key topics related to uptake of the HPV vaccine by adolescent girls.. Three caregiver discussion groups consisting of 6-8 participants were conducted at a secluded area provided within the Sub-county health facilities. The adolescent mini-FGD were conducted within school premises, consisting of 6 and 7 participants respectively. The interviews and discussions were recorded on a digital device and subsequently transcribed verbatim. On average, the surveys took 25 minutes, the KII approximately 30-45 minutes and the FGD close to an hour.

### Study variables

For this study, uptake was defined as girls within the 10-14 age bracket who had received the two recommended doses of the HPV vaccine. Having received two doses was scored as “yes” while “no” implied the converse. A four-point Likert scale was used to measure participant perceptions on inter-personal relationships, religion and culture, by querying communication related to the vaccine and the opined acceptability by family and community.

### Data management and analysis

Quantitative data were checked in the field for completeness. Survey data with no missing data were entered into Kobo tool box and then exported to Excel and SPSSv29 for descriptive and inferential statistical analysis. Bivariate analysis and Logistic regression were carried out to identify significant associations with HPV vaccine uptake. Variables returning a significant p-value (p≤0.05) were then subjected to multi-variable logistic regression analysis. Model fitness was checked using Nagelkerke *R^2^* = .28, *p* < .001. An alpha of 0.05 was considered significant for both levels of analysis. The qualitative interviews and discussions were recorded using a digital device, and transcribed verbatim. The transcripts were read and re-read, coded and arranged according to emerging sub-themes and themes, for thematic analysis and interpretation alongside quantitative findings. Excerpts from representative quotes were presented verbatim.

### Ethical considerations

Ethical clearance to conduct this study was granted by the Maseno University Scientific Ethics Review Board (MUSERC/01348/24) and research authorization from NACOSTI (P/24/37060). Written Informed Consent for participation in the study was sought from caregivers of adolescent girls, and assent obtained from the girls, prior to data collection. Informed consent was also sought from interview subjects, and all data was handled confidentially. Authorization was obtained from County and Sub-County administrators (NCCG/HWN/REC/605), as well as school administrators prior to commencement of the study. Participation was purely voluntary and participants were allowed to withdraw from the study at any time.

## Results

### Socio-Demographic Characteristics of Respondents

The Socio-Demographic Characteristics of Respondents are presented in Table 2. From the targeted 385 participants, 376 participated in the study and of these, 5 incomplete surveys were excluded from the analysis Three hundred and seventy-six girls between the ages of 10 and 14 years of age participated in the study, with a response rate of 96.36%.

**Table 2:**
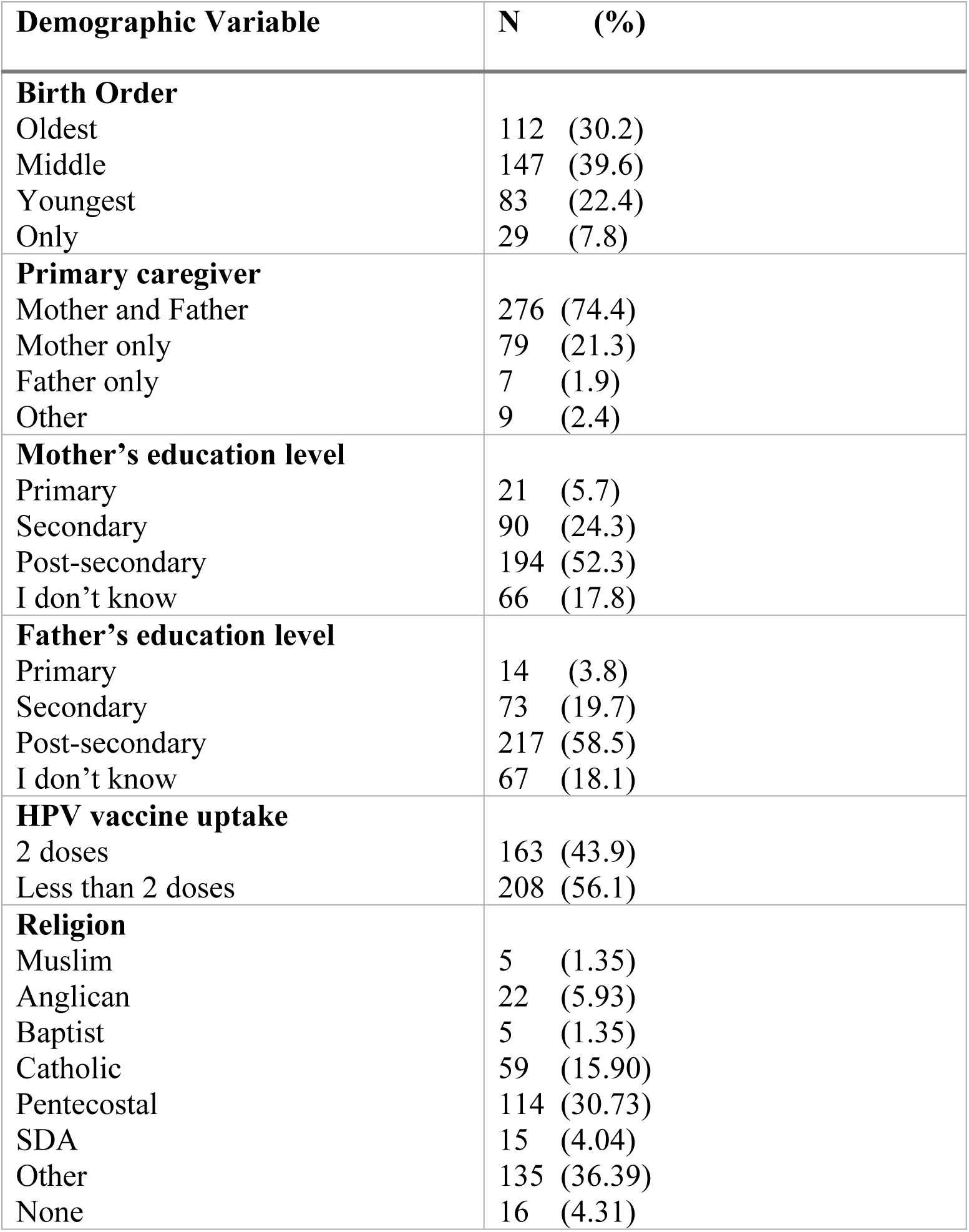
Respondent demographics.

| Demographic Variable | N | (%) |
| --- | --- | --- |

|  |  |
| --- | --- |
| **Birth Order** |  |
| Oldest | 112 (30.2) |
| Middle | 147 (39.6) |
| Youngest | 83 (22.4) |
| Only | 29 (7.8) |
| **Primary caregiver** |  |
| Mother and Father | 276 (74.4) |
| Mother only | 79 (21.3) |
| Father only | 7 (1.9) |
| Other | 9 (2.4) |
| **Mother's education level** |  |
| Primary | 21 (5.7) |
| Secondary | 90 (24.3) |
| Post-secondary | 194 (52.3) |
| I don't know | 66 (17.8) |
| **Father's education level** |  |
| Primary | 14 (3.8) |
| Secondary | 73 (19.7) |
| Post-secondary | 217 (58.5) |
| I don't know | 67 (18.1) |
| **HPV vaccine uptake** |  |
| 2 doses | 163 (43.9) |
| Less than 2 doses | 208 (56.1) |
| **Religion** |  |
| Muslim | 5 (1.35) |
| Anglican | 22 (5.93) |
| Baptist | 5 (1.35) |
| Catholic | 59 (15.90) |
| Pentecostal | 114 (30.73) |
| SDA | 15 (4.04) |
| Other | 135 (36.39) |
| None | 16 (4.31) |

Forty percent of participants had both older and younger siblings, 22.2% were the youngest and 7.8% were an only child (Table 2). Just over 23% came from single parent household, and 74.4% of respondents reported living with both parents. Over 50% of both mothers and fathers had post-secondary education, whilst 21 mothers (5.7%) and 14 fathers (3.8%) were reported to have a primary level of education. The proportion of girls who reported uptake of the vaccine (defined as two doses at least 6 months apart) was 43.9%. Conversely, 39.6% had not ever received the HPV vaccine.

Majority (96.5%) of the participating students identified as Christians, with Pentecostal church being the most common denomination, followed by Catholic while 36.8% (n=135) belonged to other smaller churches. In terms of religion, uptake was highest amongst Muslims at 60%, while Christians had the lowest uptake at 45.3%. Uptake was lowest among respondents from single-Father households (28.6%) but similar among single-Mother and dual-parent homes. Discussants in the adolescent FGDs expressed that their fathers “*had no experience*,” and “[he] *does not know those things*” as well as “*I cannot go to dad, he does not understand*.” Mothers in their FGD additionally contributed “*For many (fathers) it is because they do not know what it is or even how it helps. They just hear of it. Plus they don’t have time*” and also *“They don’t know, they have never heard from the community health promoters- they were not there.*” Another mother also offered “*You as a mom have so many secrets of your child, more than the father. You are in the house most of the time with the girl. You can take her to be injected and tell her not to say she was injected. Because if the dad has refused and you know that is something that will help her, you will take her to be injected but the father won’t know.*”

Uptake was highest for participants whose fathers had a tertiary level of education (46.5%), and mothers had a secondary level of education (48/9%). However, none of these variables were significantly associated with uptake of the HPV vaccine by our respondents.

### Familial and social relations with Adolescent girls

While 79.2% of the respondents reported being able to discuss HPV and the vaccine with their caregivers, the remaining 20.8% were either undecided or unable to (Table 3). Only 43.4% of respondents had actually had HPV-related discussions in their home, at varying lengths, but 41.5% had not discussed the topic at all. From adolescent FGD, it was stated that “*even if you talk to mom, she still has to tell dad so that they agree together.”* Half of the respondents (50.1%) believed their caregivers wanted them to be vaccinated but 22.9% believed their caregivers to be against their vaccination. This was substantiated in the adolescent FGD where a discussant said “*If a parent refuses you can’t get it*” and another “*If a parent agrees, that’s it.*” Seventy seven respondents (20.8%) affirmed having a vaccinated relative and a similar number (19.4%) reported receiving negative information about the vaccine from a relative. Among friends, 67.7% of respondents reported that their peers had a positive attitude towards the vaccine. However, only 32.1% reported they would be influenced for the vaccine by their vaccinated friends.

**Table 3:**
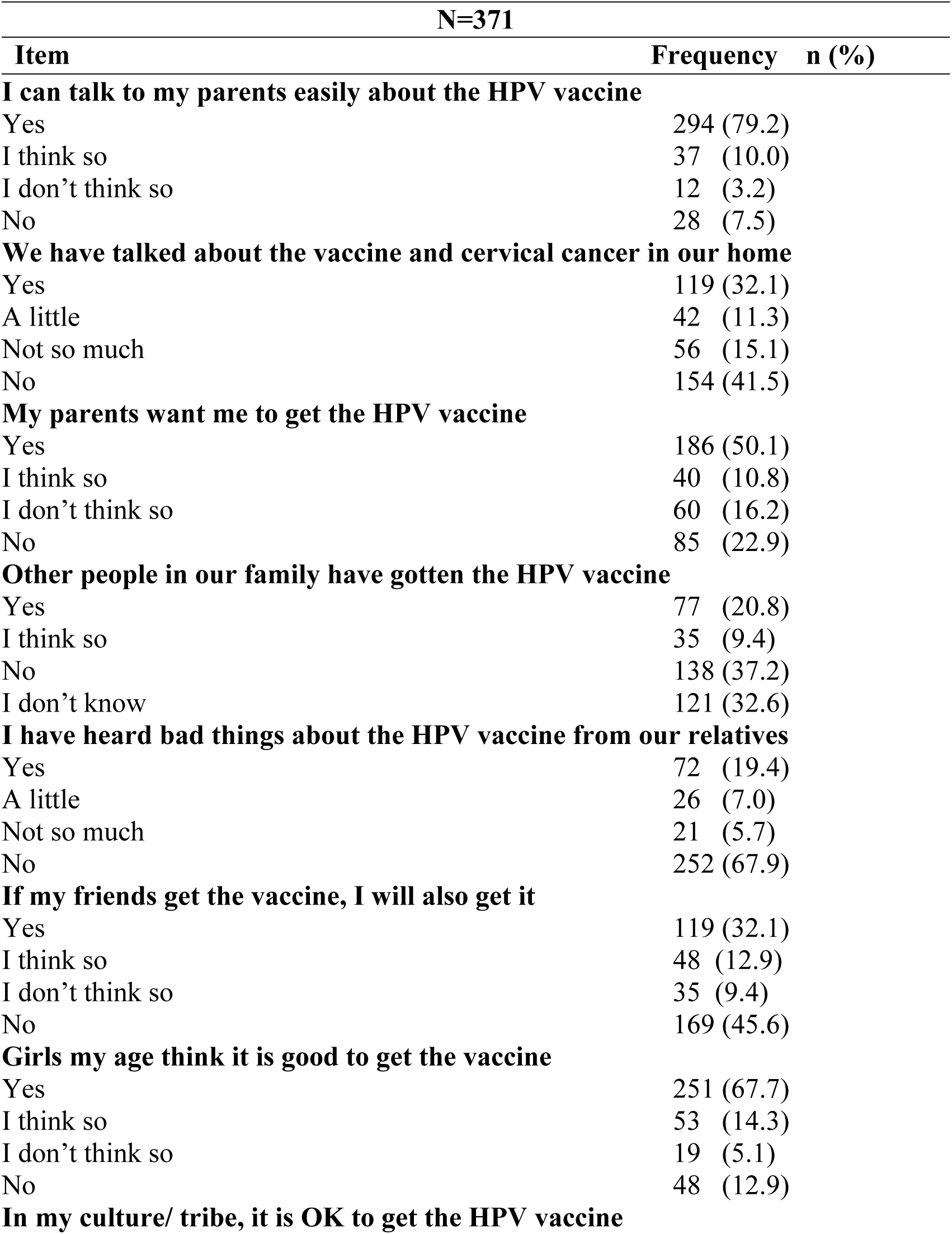
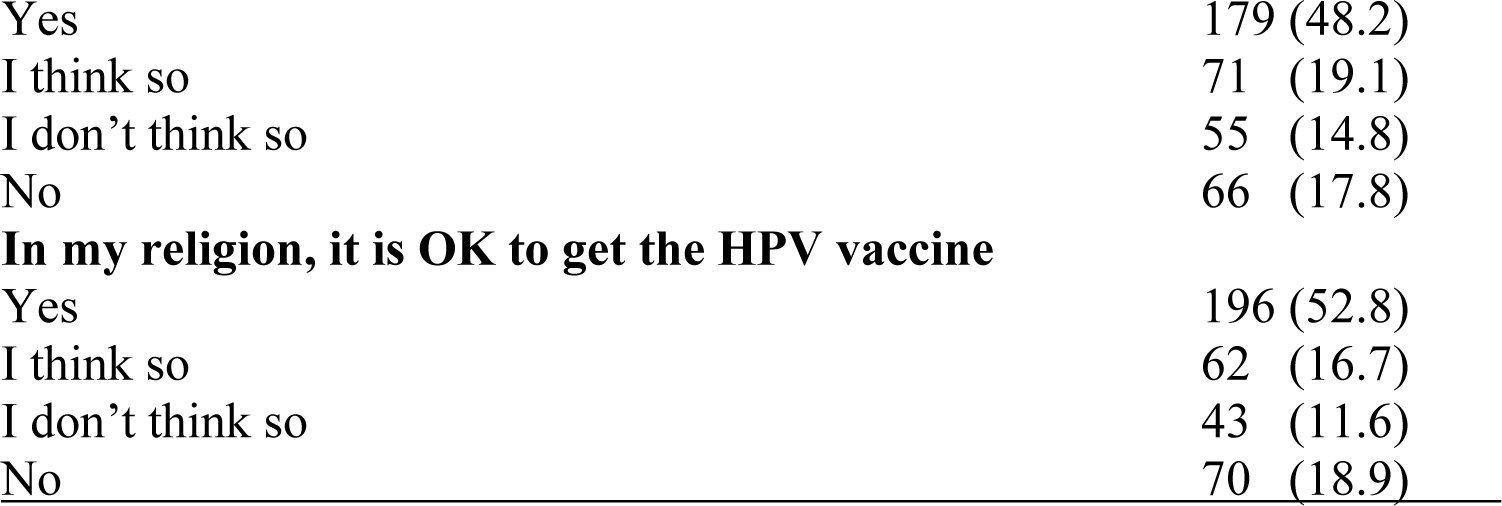
Familial and Social relations.

Uptake of the HPV vaccine was significantly associated with having talked about the vaccine in the home χ2 (3, N = 371) = 20.2, *p* < .001; parent inclination for the girl to be vaccinated χ2 (3, N = 371) = 36.8, *p* < .001; having a HPV vaccinated family member χ2 (3, N = 371) = 11.282, *p* = .01; as well as the perception of other girls towards getting vaccinated χ2 (3, N = 371) = 9.9, *p* = .02 (Table 4). However, the association between the vaccination of respondents’ peers and uptake was not significant.

**Table 4:** Association between Relationships, Religion and Culture and Vaccine Uptake.

| Item | N=371 | | $\chi^2$ | DF | P Value |
| --- | --- | --- | --- | --- | --- |
|  | Uptake<br>Yes | No |  |  |  |
| **I can talk to my parents easily about HPV and the vaccine** |  |  |  |  |  |
| Yes | 128 (43.5) | 166 (56.5) | Fisher's exact |  | 0.053 |
| I think so | 18 (48.6) | 19 (51.4) |  |  |  |
| I don't think so | 9 (75) | 3 (25) |  |  |  |
| No | 8 (28.6) | 20 (71.4) |  |  |  |
| **We have talked about the vaccine and cervical cancer in our home** |  |  |  |  |  |
| Yes | 59 (49.6) | 60 (50.4) | 20.163 | 3 | <0.001* |
| A little | 19 (45.2) | 23 (54.8) |  |  |  |
| Not so much | 36 (64.3) | 20 (35.7) |  |  |  |
| No | 49 (31.8) | 105 (68.2) |  |  |  |
| **My parents want me to get the HPV vaccine** |  |  |  |  |  |
| Yes | 110 (59.1) | 76 (40.9) | 36.770 | 3 | <0.001* |
| I think so | 13 (32.5) | 27 (67.5) |  |  |  |
| I don't think so | 13 (21.7) | 47 (78.3) |  |  |  |
| No | 27 (31.8) | 58 (68.2) |  |  |  |
| **Other people in our family have gotten the HPV vaccine** |  |  |  |  |  |
| Yes | 46 (59.7) | 31 (40.3) | 11.282 | 3 | 0.01* |
| I think so | 17 (48.6) | 18 (51.4) |  |  |  |
| No | 55 (39.9) | 83 (60.1) |  |  |  |
| I don't know | 45 (37.2) | 76 (62.8) |  |  |  |
| **I have heard bad things about the HPV vaccine from our relatives** |  |  |  |  |  |
| Yes | 35 (48.6) | 37 (51.4) | 2.362 | 3 | 0.502 |
| A little | 13 (50) | 13(50) |  |  |  |
| Not so much | 11 (52.4) | 10 (47.6) |  |  |  |
| No | 104 (41.3) | 148 (58.7) |  |  |  |
| **If my friends get the vaccine, I will also get it** |  |  |  |  |  |
| Yes | 51 (42.9) | 68 (57.1) | 5.541 | 3 | 0.137 |
| I think so | 28 (58.3) | 20 (41.7) |  |  |  |
| I don't think so | 12 (34.3) | 23 (65.7) |  |  |  |
| No | 72 (42.6) | 97 (57.4) |  |  |  |
| **Girls my age think it is good to get the vaccine** |  |  |  |  |  |
| Yes | 124 (49.4) | 127 (50.6) | 9.881 | 3 | 0.020* |
| I think so | 19 (35.8) | 34 (64.2) |  |  |  |
| I don't think so | 6 (31.6) | 13 (68.4) |  |  |  |
| No | 14 (29.2) | 34 (70.8) |  |  |  |
| **In my culture/ tribe, it is OK to get the HPV vaccine** |  |  |  |  |  |
| Yes | 99 (55.3) | 80 (44.7) |  |  |  |
| I think so | 28 (39.4) | 43 (60.6) | 20.225 | 3 | <0.001* |
| I don't think so | 18 (32.7) | 37 (67.3) |  |  |  |
| No | 18 (27.3) | 48 (72.7) |  |  |  |
| **In my religion, it is OK to get the HPV vaccine** |  |  |  |  |  |
| Yes | 100 (51.0) | 96 (49.0) |  |  |  |
| I think so | 28 (45.2) | 34 (54.8) | 12.005 | 3 | 0.007* |
| I don't think so | 12 (27.9) | 31 (72.1) |  |  |  |
| No | 23 (32.9) | 47 (67.1) |  |  |  |

\*= $P$ value significant at or below 0.05

### Influence of Religion and Culture on uptake

As many as 52.8% of respondents believed the HPV vaccine to be acceptable in their religious faith but 18.9% believed it not to be. With regard to Muslims, key informant offered *“We had foreseen there could be some resistance especially from the Muslim group…we had a consultative meeting which included Imams. We knew the other side- the Muslim side we had to engage their leader at least to speak to them in their language to understand it well so that they could be convinced that the vaccine is very safe and has got a lot of advantages for them.”*

Slightly less than half (48.2%) opined that getting the HPV vaccine would be acceptable within their cultural community, while 17.8% believed the opposite, and the rest were uncertain. Mothers from FGD explained “*You know, even if there are tribes, each parent loves their child. That is why when it comes to hospital matters, tribe is not an issue so if there is something, you are very quick to protect, as long as you understand*.” Another discussant noted that *“You find that our lack of knowledge and lack of understanding makes us fight or block some things but if you can educate people…we will understand and know the importance of it.*” A statistically significant association was found between the perceived cultural and religious acceptability of the vaccine and uptake of the HPV vaccine χ2 (3, N = 371) = 20.2, *p* < .001 and χ2 (3, N = 371) = 12.0, *p* =.007 respectively.

### Logistic regression

Binomial logistic regression performed on the significant variables failed to show significant predictive value on all but one item. (Table 5). Compared to those who had not discussed the vaccine in their homes, those who had limited conversation on the vaccine were less likely to be vaccinated.

**Table 5:**
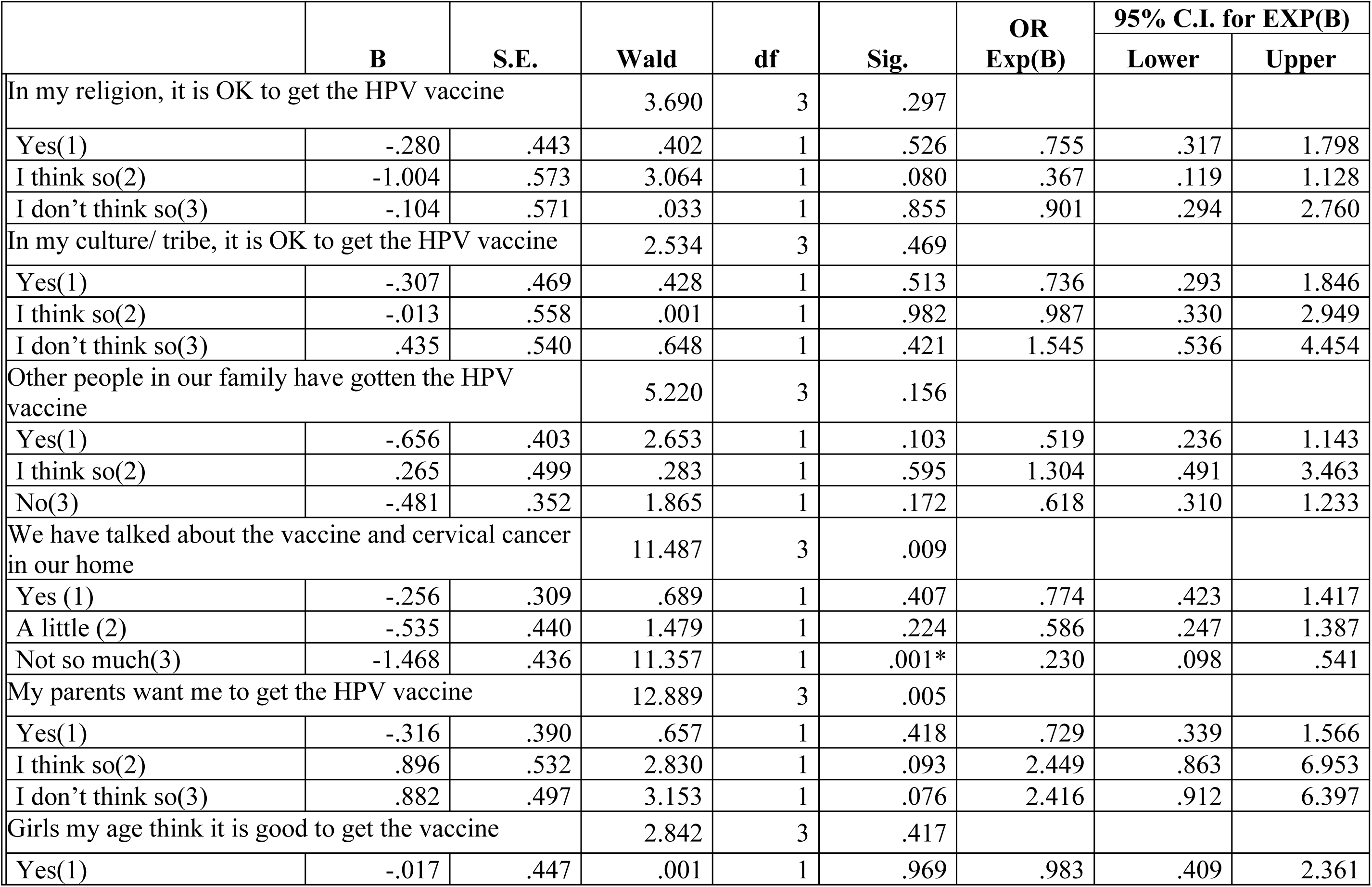
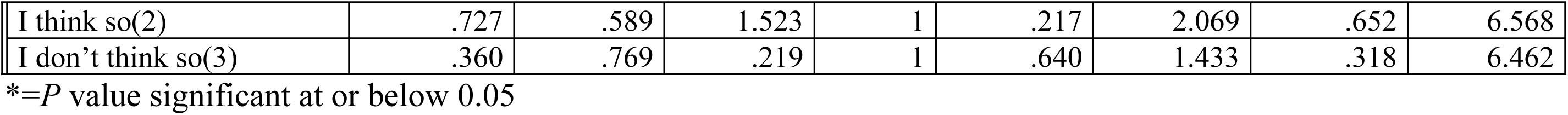
Logistic regression SPSS output.

### Discussion Demographics

The study’s reported vaccination uptake was greater (43.9%) than both the national uptake of 33 % and the county uptake level of 17.2% in 2023.The approach used, in which vaccination uptake was self-reported and may thus have been overstated, may be the cause of this discrepancy.

Furthermore, the sample of respondents came from public schools, but the county and national values encompass all females, whether they attend private or public schools or are not enrolled in school. The findings on uptake were found to be similar to studies conducted in Ethiopia among adolescent school girls (26,27) but unlike findings from a study in Uganda (28) where it was much lower at 8.6%.

The birth order of participants in this study was fairly spread out between being the youngest, middle or oldest child but was not associated with uptake. The distribution of birth order could be a reflection of the urban setting where families consist of mixed-ages, precipitated by rural-urban migration, and frequent relocation. This is unlike the findings from previous studies that report an association between birth order and uptake. For instance, Amdisen and colleagues reported lower uptake in children of higher birth order (29), while Isabirye et al concluded that a middle child was more than twice as likely to be vaccinated than a first born (30).

Many of the students resided with both parents and had similar uptake as those residing with a single female parent, which was similar to findings from a previous study where no difference on uptake was reported based on marital status (31). However, lower uptake has been reported among girls residing with one parent (29,32). The similarity in uptake between homes of single mothers and those of dual parents could suggest that mothers primarily made the decision for HPV vaccination, or heavily influenced decisions regarding the vaccine for their daughters. Respondents from single-father households reported the lowest uptake, and may allude to discomfort addressing the topic, or lack of knowledge. The attitudes of discussants in the adolescent focus groups rendered the father not only unapproachable, but also uninformed, However, studies on single Father households are lacking.

Whereas the education level of 17.8% and 18.1% of mothers and fathers respectively was reportedly unknown, over three quarters of both male and female caregivers had a secondary (high-school) or higher level of education. More educated fathers had daughters with higher uptake while mothers with high school education had higher uptake. This could be explained by heightened exposure of men with higher education, whereas high-school educated mothers may be more trusting than their college or university educated counterparts who may query more and have higher mistrust of the government or health care system. Fathers on the other hand were qualitatively reported to be generally less knowledgeable on the subject, and less available, which may explain why respondents from Single Father households also reported the lowest rate of vaccine uptake. A discussant also justified neither consulting nor informing a father and making the decision independently in her opined best interest of the girl. This concurs with the report that adolescents were less likely to be vaccinated against HPV when husbands were involved in decision making (33). A review of literature on this reveals contradictory findings on the educational level of caregivers and subsequent vaccine uptake by their daughters (34–37).

### Familial and social relationships influence on uptake of HPV

Whereas 79.2% of the respondents reported being able to openly discuss vaccine-related issues with their caregivers, slightly over half of those had actually had HPV-related discussions in their home. Additionally, 50.1% of the respondents believed their caregivers wanted them to be vaccinated. Compared to those who had not discussed the vaccine in their homes, those who had limited conversation on the vaccine were less likely to be vaccinated. This may be a result of misinformation or lack of adequate knowledge, resulting in more doubt and less confidence in the vaccine. These aspects were significantly associated with uptake of the vaccine, denoting the importance of parent-child communication for decision-making and vaccine uptake, which is also echoed in other studies (38,39). However, the decision was ultimately made by the parent as indicated by statements from adolescent FGD discussants indicating the finality of a parent’s decision. Additionally, while they more freely approached their mother, they noted that mothers still had to discuss with the fathers, hence acknowledging the father’s role in decision making. The girls in this study’s FGD were unaware of their mother’s utilization of preventative screenings. However, mothers in the FGDs confessed to having been screened for breast and cervical cancer, and to also having had their daughters vaccinated against HPV. This agrees with a finding that parents who regularly participated in health screenings and vaccination, were reportedly more likely to vaccinate their daughters against HPV (40).

The vaccination of a relative was associated with uptake of the HPV vaccine yet only a fifth (20.8%) of respondents affirmed having a vaccinated relative and a similar proportion (19.4%) reported receiving negative information about the vaccine from a relative. Family and family friends possess varying levels of influence on caregiver decision-making with regard to vaccination of their daughters against HPV drawn from health practices (41,42). The beliefs and experiences of both caregivers and relatives such as abnormal test results, actual disease or concerning family history can impact their views on the vaccine (43–46).

Additionally, two thirds of respondents reported that their peers had a positive attitude towards the vaccine, which was significant for uptake, yet only a third reported they would be influenced for the vaccine by their vaccinated friends. The effect of peer influence on vaccine uptake may have been diffused by the lack of decision-making authority. As minors, the respondents were still more likely to follow the guidance of their parents over that of their peers for health decisions. Despite peers sharing sentiments of fear and pain, seldom do adolescents have the power to decide and initiate the HPV vaccine on their own (47). Yet with adequate knowledge and support, research demonstrates the health-decision-making potential of adolescents (46–51).

### Influence of Religion and Culture on uptake

Whereas over half of respondents believed the HPV vaccine to be acceptable in their religious faith, slightly less than half considered it acceptable within their cultural community. Both perceived religious and cultural acceptability were significantly associated with uptake of the HPV vaccine among respondents. While poor uptake among Muslim faithful had been anticipated by health workers in the area, they had higher uptake than non-Muslims. This higher uptake could be explained by the urban setting, but also more importantly it could be an over-estimation due to the low number of participants who ascribed to the Muslim faith. Studies have reported lower uptake among Muslims who for instance took issue with non-halal components of the vaccine (52–54). While religion was independently significantly associated with uptake, the lack of significant predictive value could also point to additional confounding factors not addressed in the current study.

The relationship between cultural acceptability and uptake of the HPV vaccine was statistically significant, but binomial logistic regression failed to capture the strength of any association. Cultural position could therefore be surpassed by the protective value of the vaccine. As explained by mothers from FGD, the instinct to protect their child would override cultural views, especially where they had more current information. In their opinion, ignorance could cause resistance to the vaccine or health initiatives because of cultural beliefs. In literature, cultural beliefs and practices relating to household decision making patterns and sexual conduct of adolescents influenced adolescent vaccination (54,55) hence by incorporating cultural and religious nuances in community education, vaccine acceptability and uptake (56,57) would improve.

Caregiver decisions regarding the vaccine may be influenced by numerous factors including their health beliefs and experiences as well as social and cultural influences.. For instance, parents may make decisions based on their own potentially erroneous estimation of their child’s risk of exposure to the virus, guided by their religious and cultural beliefs and practices, or moral upbringing (54,58).

### Strengths and Limitations

This study employed mixed methods which allowed for triangulation of findings from quantitative results as well as various views in the qualitative data. However, because the data was drawn from school-going girls, it is difficult to generalize to all adolescent girls. Additionally, the voice of the male parent was not investigated, and would have provided valuable insights, beneficial to the HPV vaccination programs.

## Conclusion

The findings of the present study suggest the importance of family values in the medical decision-making process of caregivers for their children. Community-wide educational efforts that target male and female caregivers and adolescents while taking into account cultural and religious spheres would encourage open communication, enhance understanding and dispel myths around HPV and the vaccine. Given the strong association between caregiver-child dialogue and uptake, interventions that equip families, build confidence, and facilitate such dialogue could be used to drive uptake of the vaccine. Additional studies are needed to examine health decisions in households of single fathers parenting girls.

S1 FGD Guide for Girls

S2 FGD Guide for Caregivers

S3 Regression Analysis

## Data Availability

All relevant data are within the manuscript and its Supporting Information files.

